# EFFECT OF HEALTH EDUCATION IN THE CONTROL OF SCHISTOSOMIASIS IN DASS EMIRATE COUNCIL OF BAUCHI STATE, NIGERIA: AN INTERVENTION STUDY

**DOI:** 10.1101/2024.07.11.24310273

**Authors:** Sunday Charles Adeyemo, Gbadebo Jimoh Oyedeji, James Atolagbe, Oladunni Opeyemi, Sunday Olanrewaju, Calistus Akinleye, Funsho Olagunju, Eniola Dorcas Olabode, Abdulwaris Salisu Maleka

## Abstract

**Objective:** Schistosomiasis has been recognized by WHO as major contributor to mortality and morbidity particularly in Sub Sahara Africa where it is mostly prevalent. There is a lack of reliable data on the effectiveness of health education interventions in reducing the prevalence of Schistosomiasis in Bauchi State. Hence, the study assessed the prevalence of Schistosomiasis and the knowledge, attitude and practices of community members of Dass Emirate towards the prevention and control of Schistosomiasis before and after health education intervention.

**Results:** At pre-intervention, majority of the respondents 234 (66.9%) have been diagnosed, or have family members or community members who have been diagnosed with Schistosomiasis. Ninety-six (27.5%) of respondents have good knowledge about Schistosomiasis. 79 (22.6%) of the respondents strongly agree that they can confidently recognize symptoms of schistosomiasis. Only 91 (26.0%) strongly agreed to taking responsibilities for taking preventive measures. At post-intervention, the prevalence of Schistosomiasis dropped to 55.1%. This was statistically significant at p =0.043. Knowledge about Schistosomiasis increased from 27.5% to 87.0% at post-intervention. This was statistically significant at p <0.05. Regarding attitude and practices, good attitude and practices increased from 59.1% at pre-intervention 71.0% at post-intervention. However, this was not statistically significant (p>0.05).

## Introduction

Schistosomiasis, commonly known as bilharzia, is a parasitic disease caused by trematode flatworms of Schistosoma. It is a major public health concern in many tropical and subtropical regions, affecting millions worldwide. The disease is transmitted through contact with fresh water contaminated with parasitic larvae, which penetrate human skin and mature in the blood vessels, leading to various clinical manifestations. [1] Globally, schistosomiasis is recognized as a significant contributor to morbidity and mortality, especially in sub-Saharan Africa. The World Health Organization (WHO) estimates that over 200 million people are infected with schistosomiasis worldwide, with most cases occurring in Africa. [2] The disease can lead to severe health complications, including liver and spleen enlargement, kidney damage, bladder cancer, and even death if left untreated. [3]

In a study on the challenges of controlling schistosomiasis through mass drug administration (MDA) campaigns were highlighted. While MDA has shown short-term benefits in reducing the prevalence and intensity of infection, the long-term impact remains uncertain. [4] The study emphasized the need for comprehensive strategies, including health education and improved sanitation, to achieve sustainable disease control. There is a lack of reliable data on the effectiveness of health education interventions in reducing the prevalence of Schistosomiasis in Bauchi State. This knowledge gap makes it difficult to design effective control programs and allocate resources efficiently. Therefore, this study aims to evaluate the effectiveness of health education as an intervention in controlling Schistosomiasis in Dass Emirate Council of Bauchi State.

## Methods

The study employed a quasi-experimental design which involved pre-intervention data collection, intervention and post-intervention data collection. The intervention was a health education intervention programme which was designed with training manual adapted from Federal Ministry of Health manual. The training targeted major stakeholders including school pupils, market people, religious leaders, farmers and herders in 10 communities of Dass Emirate of Bauchi where schistosomiasis are mostly prevalent. Self-administered semi-structured questionnaire was used to collect baseline data and post intervention data. The Fisher’s formula (n=z^2^pq/d^2^) was used to estimate a total of 450 respondents. Multistage sampling technique was used to recruit respondents. The study population of this study encompassed residents of the Dass Emirate Council. Thed data collected were analyzed with SPSS software package version 25 and p - value less than 0.05 was taken as statistically significant. Descriptive statistics were done for all variables. Comparison of variables at pre- and post-intervention was done using chi-square as a test of association.

## Results

At pre-intervention, Majority of the respondents 234 (66.9%) have been diagnosed, or have family members or community members who have been diagnosed with Schistosomiasis. Ninety-five (27.1%) reported that Schistosomiasis is very common in their community, 131 (37.5%) reported that Schistosomiasis is somewhat common in their community, 43 (12.3%) reported that Schistosomiasis is rare in their community while 81 (23.1%) are unsure about the commonness of Schistosomiasis in their community. More than half, (207, 59.1%) reported that someone in their community has been diagnosed with Schistosomiasis.

About knowledge of Schistosomiasis at pre-intervention, overall good knowledge was 27.5%. Majority (310, 88.6%) of the respondents have heard about Schistosomiasis. Majority of the respondents 200 (64.5%) heard about Schistosomiasis from health care workers. One hundred and twenty-eight (41.3%) of the respondents reported abdominal pain as a common symptom of Schistosomiasis, 36 (11.6%) respondents reported blood in urine or stool as a common symptom of Schistosomiasis, 71 (22.9%) reported fatigue as a common symptom of Schistosomiasis, while 85 (27.4%) respondents reported weight loss as a common symptom of Schistosomiasis. About transmission of Schistosomiasis, 83 (26.8%) reported that it can be transmitted through infected water sources, 140 (45.2%) reported that it can be transmitted through mosquito bites, 75 (24.2%) reported that it can be transmitted through contaminated food while 52 (16.8%) reported that it can be transmitted through human contact. Ninety-one (29.4%) reported that Schistosomiasis is preventable, while 140 (45.2%) reported that it is not preventable and 79 (25.4%) do not know. On the preventive measures of Schistosomiasis, out of 91 that said it can be prevented, 20 (30.0%) reported that it can be prevented by avoiding contact with contaminated water, 31 (34.1%) reported that it can be prevented by boiling or treating water before use, 29 (31.9%) reported that it can be prevented by proper sanitation and hygiene practices while 10 (11.0%) reported that it can be prevented by use of insecticide-treated nets.

About attitude and practices towards the prevention of schistosomiasis, at pre-intervention, 79 (22.6%) of the respondents strongly agree that they can confidently recognize symptoms of schistosomiasis. Fifty-five (15.7%) strongly agree that there is community engagement in efforts towards prevention of Schistosomiasis. Only 91 (26.0%) strongly agreed to taking responsibilities for taking preventive measures and only 23 (6.6%) strongly agreed that they discuss prevention of Schistosomiasis with their family. However, 118 (33.7%) agreed that their actions can reduce the risk of Schistosomiasis. One hundred and twenty (34.3%) avoided contact with contaminated water to prevent and control Schistosomiasis, 51 (14.6%) boiled or treated water to prevent and control Schistosomiasis, 129 (36.8%) practiced proper hygiene and sanitation to prevent and control Schistosomiasis while 50 (14.3%) used insect treated nets to prevent and control Schistosomiasis.

At post-intervention, the prevalence reduced to 190 (55.1%) and this was statistically significant at p=0.043. (Figure 1) Knowledge of schistosomiasis increased to 87.0% at post-intervention and this was statistically significant at p <0.001. (Figure 2) About attitude and practices towards schistosomiasis prevention and control, 126 (36.5%) of the respondents strongly agree that they can confidently recognize symptoms of schistosomiasis. This difference was not statistically significant at p=0.045. Eighty-eight (25.5%) strongly agree that there is community engagement in efforts towards prevention of Schistosomiasis. However, this difference was not statistically significant (p>0.05). One hundred and eighteen (34.2%) strongly agreed to taking responsibilities for taking preventive measure. The difference is statistically significant at p = 0.035. One hundred and fifty-eight (45.8%) agreed that they discuss prevention of Schistosomiasis with their family. This difference was statistically significant at p=0.022. One hundred and twenty-four (35.9%) respondents strongly agreed that their actions can reduce the risk of Schistosomiasis. However, this was not statistically significant (p>0.05). About actions taken to prevent schistosomiasis, 120 (34.8%) reported avoiding contact with contaminated water, 51 (14.8%) reported boiling or treating water before use, 124 (35.9%) reported proper sanitation and hygiene practices and 50 (14.5%) reported use of insecticide-treated nets. However, this difference was not statistically significant (p>0.05) (Table 1)

**Fig. 1.**
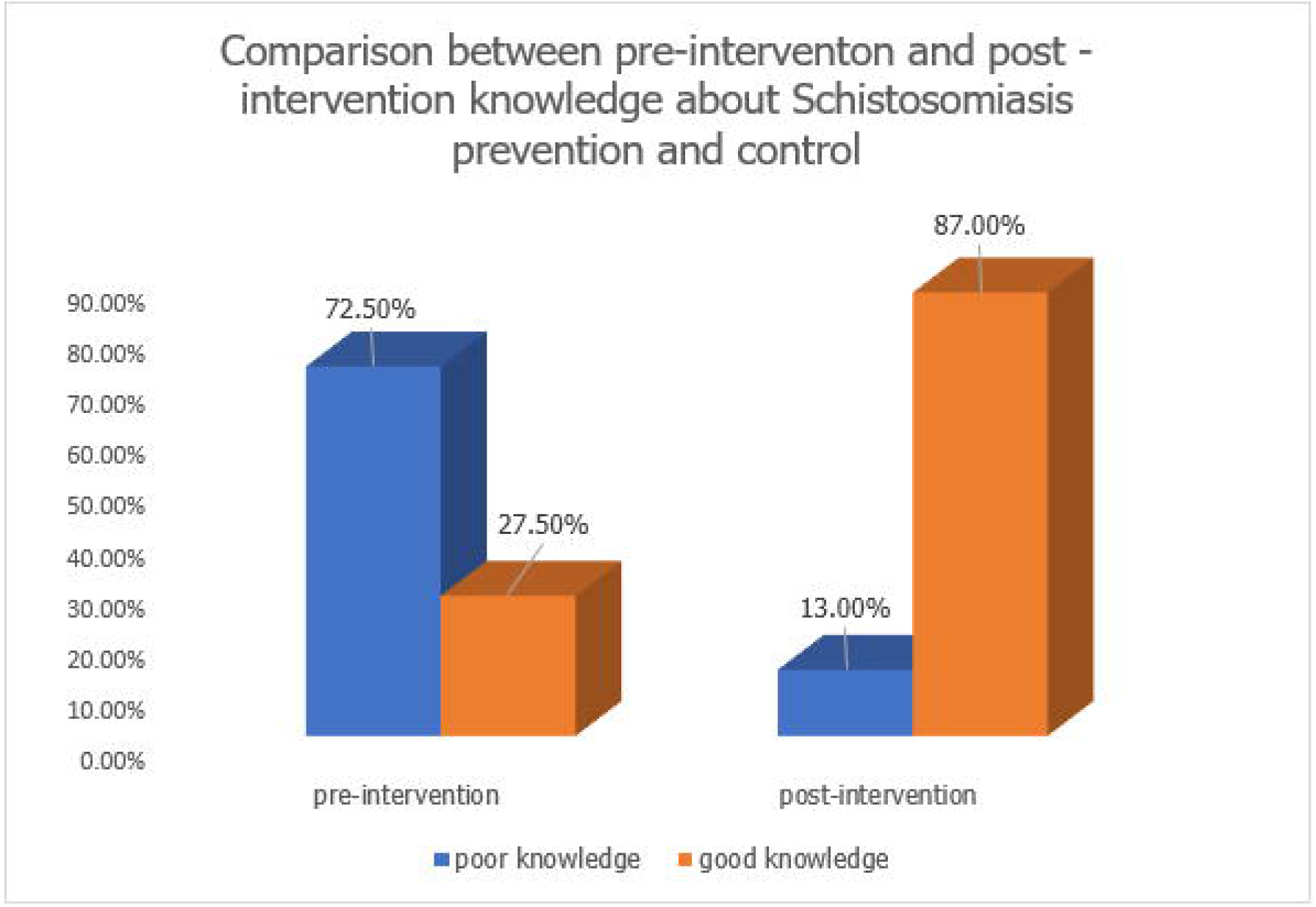

**Fig. 2.**
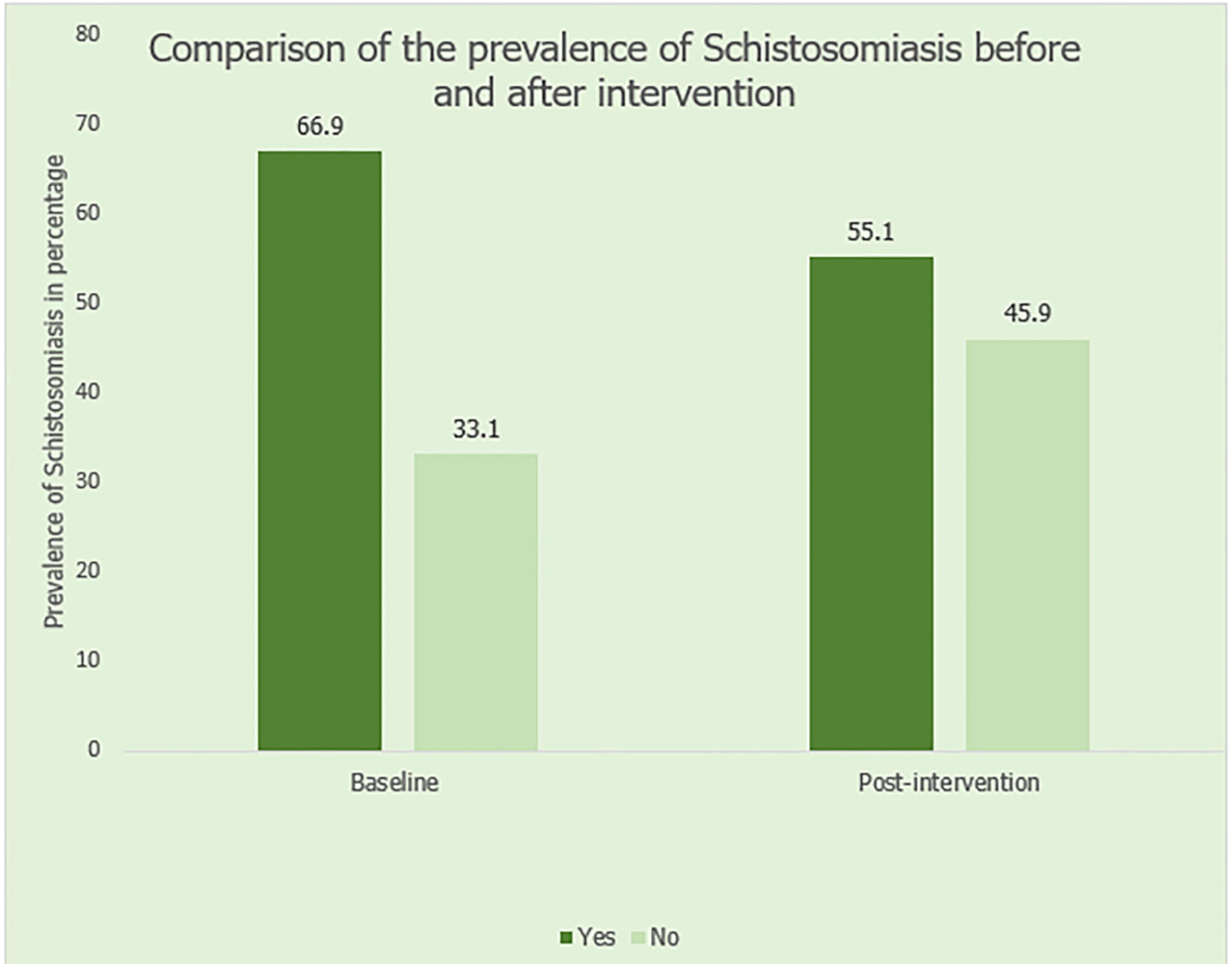

**Table 1:** Comparison of Attitude and Practices towards Schistosomiasis prevention and control among respondents at pre-intervention and post-intervention.

| Attitude and practices towards prevention and control of Schistosomiasis | Frequency (%) |  | Statistics |
| --- | --- | --- | --- |
|  | Baseline (n=350) | Post-intervention (n=345) |  |
| **I can confidently recognize Schistosomiasis symptoms** | | | $\chi^2 = 2.242$<br>df = 5<br>**p = 0.045\*** |
| Strongly Disagree | 109 (31.1) | 50 (20.4) |  |
| Disagree | 71 (20.3) | 51 (14.8) |  |
| Neutral | 55 (15.7) | 49 (14.3) |  |
| Agree | 36 (10.3) | 69 (20.0) |  |
| Strongly agree | 79 (22.6) | 126 (36.5) |  |
| **There is community engagement in prevention efforts** | | | $\chi^2 = 13.698$<br>df = 5<br>p = 0.058 |
| Strongly disagree | 120 (34.3) | 59 (17.1) |  |
| Disagree | 91 (26.0) | 64 (18.6) |  |
| Neutral | 49 (14.0) | 48 (13.9) |  |
| Agree | 35 (10.0) | 86 (24.9) |  |
| Strongly agree | 55 (15.7) | 88 (25.5) |  |
| **I take responsibility for taking preventive measures** | | | $\chi^2 = 15.090$<br>df = 5<br>**p = 0.035\*** |
| Strongly disagree | 108 (30.9) | 20 (5.8) |  |
| Disagree | 55 (15.7) | 56 (16.2) |  |
| Neutral | 37 (10.5) | 47 (13.6) |  |
| Agree | 59 (16.9) | 104 (30.1) |  |
| Strongly agree | 91 (26.0) | 118 (34.2) |  |
| **I have discussions about prevention with family** | | | $\chi^2 = 12.781$<br>df = 5<br>**p = 0.022\*** |
| Strongly disagree | 141 (40.2) | 54 (15.7) |  |
| Disagree | 70 (20.0) | 52 (15.1) |  |
| Neutral | 65 (18.6) | 17 (4.9) |  |
| Agree | 51 (14.6) | 158 (45.8) |  |
| Strongly agree | 23 (6.6) | 64 (18.6) |  |
| **I believe that my actions can reduce risk of Schistosomiasis** | | | $\chi^2 = 18.889$<br>df = 5<br>p = 0.055 |
| Strongly disagree | 90 (25.7) | 51 (14.8) |  |
| Disagree | 45 (12.9) | 53 (15.4) |  |
| Neutral | 36 (10.3) | 40 (11.6) |  |
| Agree | 118 (33.7) | 77 (22.3) |  |
| Strongly agree | 61 (17.4) | 124 (35.9) |  |
| **Actions you/ family members has taken to prevent Schistosomiasis** | | | $\chi^2 = 18.889$<br>df = 4<br>p = 0.075 |
| Avoiding contact with contaminated water | 80 (22.9) | 120 (34.8) |  |
| Boiling or treating water before use | 81 (23.1) | 51 (14.8) |  |
| Proper sanitation and hygiene practices | 89 (25.4) | 124 (35.9) |  |
| Use of insecticide-treated nets | 100 (28.6) | 50 (14.5) |  |

$\chi^2$ = Chi-square, df= degree of freedom, p= p-value, \*= statistically significant

## Discussion

The prevalence of Schistosomiasis was high at baseline. More than 3 out of 5 respondents have been diagnosed with Schistosomiasis. This can be attributed to the fact that two-fifth of the respondents are adolescents and are prone to activities that expose them to the disease. Also, more than four-fifth are rural dwellers which are prone to poor sanitation and hygiene practices. This is similar to a study who reported that children and adolescents are more prone to Schistosomiasis and poor sanitation is also linked with the disease. [5]

The Knowledge regarding Schistosomiasis was poor at baseline. Only about one-third had good knowledge about Schistosomiasis. More than four-fifth of the respondents have heard about Schistosomiasis and two-fifth of the respondents reported abdominal pain as a common symptom of Schistosomiasis. More than two-fifth reported that it can be transmitted through mosquito bites. One-third reported that Schistosomiasis is preventable, out of which one-third reported that it can be prevented by proper sanitation and hygiene practices. The poor knowledge can be attributed to low level of education among respondents. This is similar to the findings on the knowledge attitude and practices regarding rural communities in Kano State. [6]

The attitude and practices towards Schistosomiasis prevention and control was poor at baseline. One-third of the respondents reported that there was no community engagement in efforts towards prevention of Schistosomiasis. Less than one-third were taking responsibilities for taking preventive measures and only less than one-tenth of the respondents discuss prevention of Schistosomiasis with their family. However, slightly above one-third of the respondents agreed that some actions can reduce the risk of Schistosomiasis. In order to prevent and control schistosomiasis, slightly above one-third avoided contact with contaminated water and practiced proper hygiene and sanitation. This reveals significant shifts in the community’s perceptions and reported experiences with the disease.

At post-intervention, the prevalence of Schistosomiasis decreased significantly to slightly more than half, which was statistically significant at p=0.043. This could be attributed to heightened community awareness of Schistosomiasis as a result of health education, which aligns with research that suggests heightened awareness through health education can increase community concern thereby reducing prevalence of Schistosomiasis. [7]

At post-intervention, overall knowledge improved from less than one-third at baseline to fourth-fifth after health education intervention. This aligns with the findings which reported that improved symptom recognition was linked to effective health education, potentially leading to earlier disease detection and treatment. [8] The attitude and practices concerning Schistosomiasis control improved after the health education intervention. One-third of the respondents strongly agreed to taking responsibilities for taking preventive measures while more than two-fifth of the respondents agreed that they discuss prevention of Schistosomiasis with their family. A study by [9] supports this finding, suggesting that enhanced community engagement is often observed following targeted health education campaigns, which empower communities to take an active role in disease prevention. The role of family discussion in promoting health practices has also been well-documented, [7] and the result of this study echo the potential of family-based interventions to amplify health messages within a community. More than one-third of the respondents strongly agreed that some actions can reduce the risk of Schistosomiasis. This is indicative of an internalization of health education messages, where community members not only learn about preventive measures but also believe in their efficacy. This internal belief system is essential for sustained behavior change and has been cited in previous studies as a critical component of health education success. [10] The increase in the items indicating Knowledge, Attitude and Practices may be attributed to an enhancement in disease detection and diagnosis efforts, a finding consistent with studies, that demonstrate improved identification of endemic diseases following concerted health interventions. [11] Similar patterns have also been noted in other literature, where increased awareness following educational interventions correlates with improved health behaviors. [12]

An essential aspect of the study’s success was its culturally sensitive approach. By tailoring the intervention to fit the local cultural context and incorporating local beliefs and practices, the program achieved higher engagement and effectiveness. This approach is in line with the recommendations of [7], which emphasize the importance of cultural sensitivity in health education interventions.

The findings highlight the importance of health education interventions in improving knowledge, attitudes, and practices towards Schistosomiasis control. They underscore the need for addressing perceived barriers and emphasize the role of community engagement in Schistosomiasis prevention and control. The study recommends prioritizing formulation of comprehensive health policies that address Schistosomiasis within the broader context of public health and increased allocation of resources while encouraging partnership from stakeholders and participation from individuals in combatting schistosomiasis.

## Data Availability

All data produced in the present study are available upon reasonable request to the authors

## Acknowledgements

The authors wish to acknowledge the respondents and the spouses of the authors for their understanding during the course of this study.

## Limitation

The limitation of the study is the reliance on self-reported data, which may be subject to recall bias and social desirability bias. Future research could consider using objective measures, such as medical records, to validate self-reported prevalence of Schistosomiasis. The duration of the health education was also limited due to the nature of occupation of the respondents who were farmers who needed to go to their various farms and the insecurity issues peculiar with the study location.

## Declarations

### Ethical approval

Ethical approval was obtained from Research Ethics Committee of Ministry of Health, Bauchi State and Research Ethics Committee of Adeleke University, Ede. Informed consent was also obtained from participants after providing them with detailed information about the study’s purpose, the procedures involved, the potential risks and benefits, and their rights as participants. Confidentiality was strictly maintained throughout the study. All participant data were anonymized and stored securely, accessible only to the research team. Identifiers were removed or coded to prevent the identification of individual participants in any reports or publications resulting from the research.

### Consent for publication

Not applicable

### Availability of data and material

The data for this study is available on reasonable request from the corresponding author.

### Authors’ contribution

Sunday Charles Adeyemo-Study design, data collection and analysis, Gbadebo Jimoh Oyedeji-Study design and training of health educators, James Atolagbe-Supervision, Oladunni Opeyemi-Study design and supervision, Sunday Olanrewaju-Ethical compliance, Calistus Akinleye-Study design, Funsho Olagunju-Data analysis, Eniola Dorcas Olabode-Data Analysis and manuscription, Abdulwaris Salisu Maleka-Data collection

